# Modelling the impact of rapid tests, tracing and distancing in lower-income countries suggest that optimal policies vary with rural-urban settings

**DOI:** 10.1101/2021.03.17.21253853

**Authors:** Xilin Jiang, Wenfeng Gong, Zlatina Dobreva, Ya Gao, Matthew Quaife, Christophe Fraser, Chris Holmes

## Abstract

Low- and middle-income countries (LMICs) remain of high potential for hotspots for COVID-19 deaths and emerging variants given the inequality of vaccine distribution and their vulnerable healthcare systems. We aim to evaluate containment strategies that are sustainable and effective for LMICs. We constructed synthetic populations with varying contact and household structures to capture LMIC demographic characteristics that vary across communities. Using an agent-based model, we explored the optimal containment strategies for rural and urban communities by designing and simulating setting-specific strategies that deploy rapid diagnostic tests, symptom screening, contact tracing and physical distancing. In low-density rural communities, we found implementing either high quality (sensitivity > 50%) antigen rapid diagnostic tests or moderate physical distancing could contain the transmission. In urban communities, we demonstrated that both physical distancing and case finding are essential for containing COVID-19 (average infection rate < 10%). In high density communities that resemble slums and squatter settlements, physical distancing is less effective compared to rural and urban communities. Lastly, we demonstrated contact tracing is essential for effective containment. Our findings suggested that rapid diagnostic tests could be prioritised for control and monitor COVID-19 transmission and highlighted that contact survey data could guide strategy design to save resources for LMICs. An accompanying open source R package is available for simulating COVID-19 transmission based on contact network models.

## Introduction

The severe acute respiratory syndrome coronavirus 2 (SARS-CoV-2), which causes the COVID- 19, has rapidly spread worldwide since the end of 2019. ^1^ Inadequate vaccine distribution and insufficient healthcare resources have contributed to many low- and middle- income countries (LMICs) suffering more from the pandemic than affluent countries. ^2^ Many LMICs rely on testing, tracing and physical distancing to control and monitor COVID-19, ^3,4^ such as test, travel restriction, and testing and tracing programs, given the inequitable distribution of vaccines and medication. While contributing to COVID-19 control and surveillance, these non-pharmaceutical interventions (NPIs) may cause considerable damage to the economies as people put their life and work on hold. ^5^ Therefore, identifying effective NPIs for LMICs could prepare the government for new variants and peaks while easing the burden on the economy and society. ^4^

Emerging immunity-escaping variants from LMICs alert us to the importance of consistent surveillance and containment of COVID-19. ^6,7^ Affordable and effective testing methods are crucial to reduce the economic burden of LMICs. Many COVID-19 containment policies rely on the real-time reverse transcription-polymerase chain reaction (RT-PCR), which can be challenging to implement in LMIC because of the limited clinical resources ^8,9^ and laboratory capacities. ^10,11^ The financial and lab personnel cost of maintaining large scale PCR testing would put a heavy burden on the vulnerable healthcare systems of LMICs. Therefore, it is essential to leverage alternative testing methods, such as rapid diagnostic tests (RDT) based on antigen detection, ^12–14^ which is cheaper and less resource-demanding. Additionally, community healthcare workers could perform contact tracing and quarantine to further mitigate the SARS-CoV-2 transmission, which has been implemented successfully during HIV and Tuberculosis outbreaks. ^15^ Combining RDT, contact tracing and quarantine, we can increase the testing capacity, ensure early and accurate detection of infected cases, contain disease transmission, and reduce the speed of viral mutation accumulation.

In this study, we evaluated the dynamics of COVID-19 transmission in younger populations that are typical of LMICs and the effectiveness of testing, tracing and distancing. We constructed synthetic contact networks using contact numbers and demographic structures in lower-income communities. To capture the different infection rates across geographical locations, ^16,17^, we simulated SARS-CoV-2 transmission in various contact structures and captured the uncertainty of the epidemic sizes using bootstrap samples of contact networks. We evaluated the realistic combined strategy involving test and tracing using PCR and RDT, advising patients to self isolate, and physical distancing. Our simulation results suggest the level of containment required to contain COVID-19 depends on contact frequency in the communities. We also showed that antigen RDT and symptom-based diagnosis could be deployed in several settings for better containment outcomes. To help researchers replicate results with demographics that are not included in our analysis and to update the results using properties of new variants, we released an open-source R package for contact network based transmission simulation which accommodates customised parameter settings.

## Material and methods

### Processing demographic data

We cited the age distribution and household sizes from the United nation. ^18,19^ (Supplementary Table 1) We considered three age groups (0-14, 15-25 and 25+ years old) for simplicity and to capture the susceptibility of the younger population and contact structure differences between younger and older groups. We cited the contact survey data from a study in Uganda to compute age-specific contact matrices and contact distribution within households.^20^ To compare geographies that have different contact numbers, we simulated contact networks that correspond to LMICs using PERC survey data from Africa CDC. ^21^ When using the contact matrices in simulation, we scaled the contact matrix to match the average daily contact number under each setting. (Table 1)

**Table 1:**
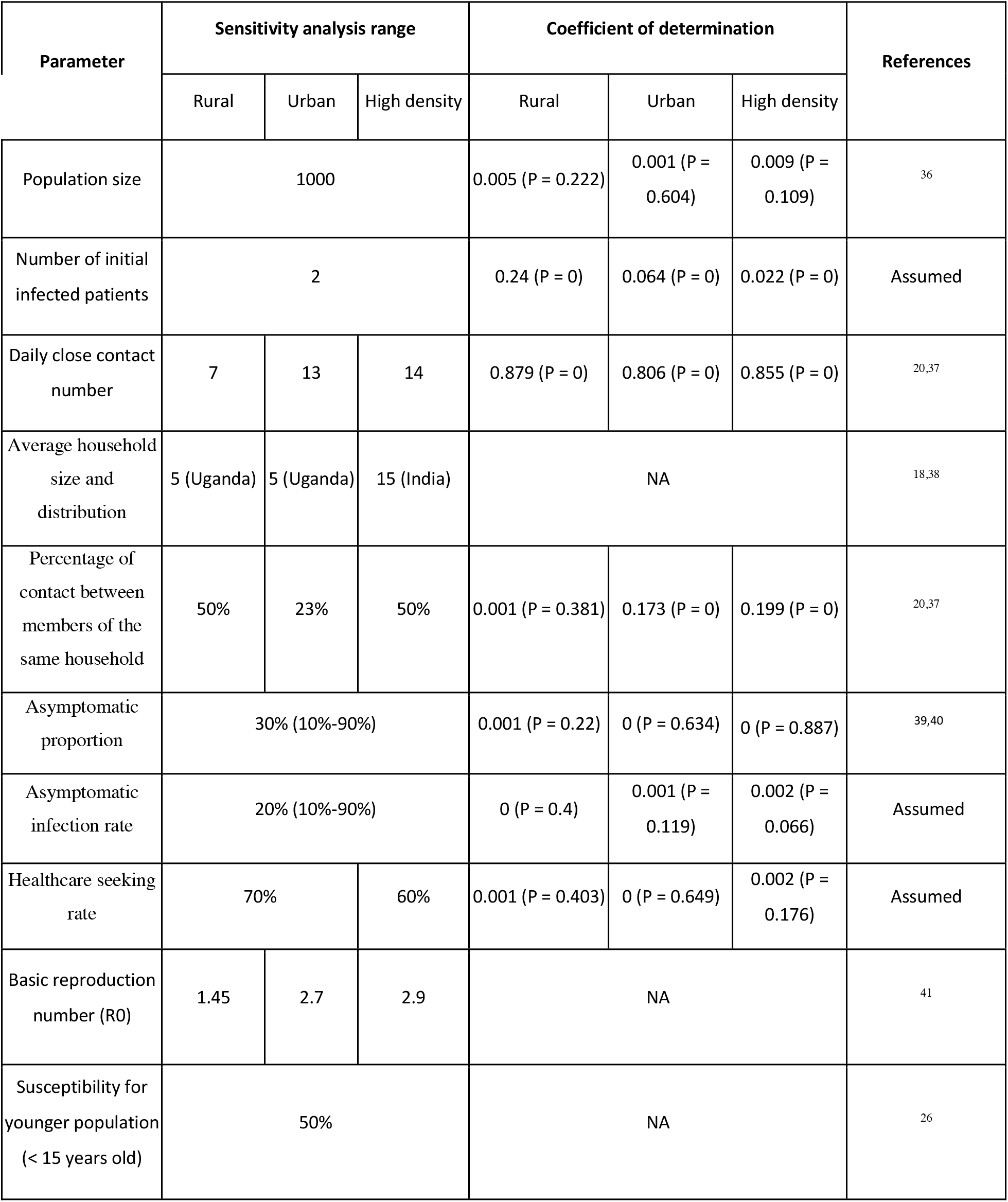
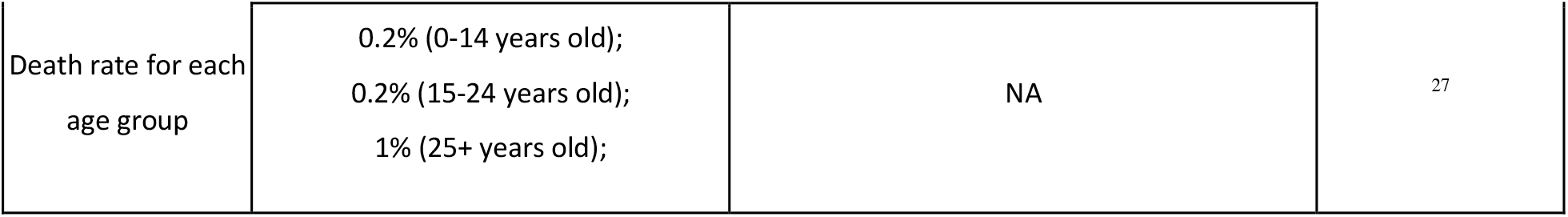
Summary of population characteristics and transmission parameters and their impact on transmission. For parameters where the source is not available for at least one of the three community settings, we simulated models with different choices of values and used regression to evaluate their impact on model outputs. (Supplementary Table 4, Supplementary Methods Section 4) The coefficient of determination (R^2^) shows the proportion of variance in the infected number that is captured by the parameter; The P-values in the parentheses show the rate of type one error of rejecting the null hypothesis that the parameter is not correlated with infection number. Last column shows the sources for cited parameters.

| Parameter | Sensitivity analysis range |  |  | Coefficient of determination |  |  | References |
| --- | --- | --- | --- | --- | --- | --- | --- |
|  | Rural | Urban | High density | Rural | Urban | High density |  |
| Population size | 1000 |  |  | 0.005 (P = 0.222) | 0.001 (P = 0.604) | 0.009 (P = 0.109) | 36 |
| Number of initial infected patients | 2 |  |  | 0.24 (P = 0) | 0.064 (P = 0) | 0.022 (P = 0) | Assumed |
| Daily close contact number | 7 | 13 | 14 | 0.879 (P = 0) | 0.806 (P = 0) | 0.855 (P = 0) | 20,37 |
| Average household size and distribution | 5 (Uganda) | 5 (Uganda) | 15 (India) | NA |  |  | 18,38 |
| Percentage of contact between members of the same household | 50% | 23% | 50% | 0.001 (P = 0.381) | 0.173 (P = 0) | 0.199 (P = 0) | 20,37 |
| Asymptomatic proportion | 30% (10%-90%) |  |  | 0.001 (P = 0.22) | 0 (P = 0.634) | 0 (P = 0.887) | 39,40 |
| Asymptomatic infection rate | 20% (10%-90%) |  |  | 0 (P = 0.4) | 0.001 (P = 0.119) | 0.002 (P = 0.066) | Assumed |
| Healthcare seeking rate | 70% |  | 60% | 0.001 (P = 0.403) | 0 (P = 0.649) | 0.002 (P = 0.176) | Assumed |
| Basic reproduction number (R0) | 1.45 | 2.7 | 2.9 | NA |  |  | 41 |
| Susceptibility for younger population (< 15 years old) | 50% |  |  | NA |  |  | 26 |

|  |  |  |  |
| --- | --- | --- | --- |
| Death rate for each age group | 0.2% (0-14 years old);<br>0.2% (15-24 years old);<br>1% (25+ years old); | NA | 27 |

### Agent-based modelling

We used an agent-based model and configured the communities based on the age distributions and household sizes cited from a Uganda study. ^20^ We simulated populations using contact numbers that span a range of African countries. Additionally, we abstracted three representative community settings that have distinct demographics: (1) a rural community which represent geographies with low contact rates and medium household sizes; (2) an urban community which represent geographies with high contact rates and medium household sizes; (3) a high density community which represent high contact number and large household sizes, which aim to capture communities such as slums and squatter settlements. (Table 1) For each setting, we simulated 20 synthetic populations, where each household was composed of at least one adult above age 25, and each household was assigned to a location on a two-dimensional plane. The contact networks for the synthetic population were configured for the corresponding household structure, age-specific contact matrices, and geographical clustering, using an Exponential Random Graph Model (ERGM). ^22^ See Supplemental Methods for details on the ERGM.

After constructing the synthetic populations with contact and household structure, we randomly choose one of the 20 synthetic populations and simulated COVID-19 transmission initiated by importing 2 cases. At each time step of a simulated outbreak, one realised contact network of ERGM was sampled to represent the daily dynamic of contacts. (Figure 1) Infection events were sampled among the contacts proportional to the transmissibility multiplied by generation time, which is a Weibull distribution that has a mode on the day of onset. ^23^ The day of onset for each infected individual was sampled from the cited incubation time distribution ^24^. The susceptibility of the younger group (0-14 years old) was set to 50% of that of the older group (15+ years old). ^25,26^ The death rate is set as 1% for the older group (25+ years old) and 0.2% for the younger group (0-25 years old). ^27^ Using the age-dependent susceptibility and basic reproduction number, we computed the next generation matrix and the transmissibility. (Supplementary Methods) At each infection event, we sampled whether an infected individual is asymptomatic using a Bernoulli distribution of probability equal to 30%. (Table 1) For detailed parameterization of transmission dynamics, see Supplementary Methods.

**Figure 1:**
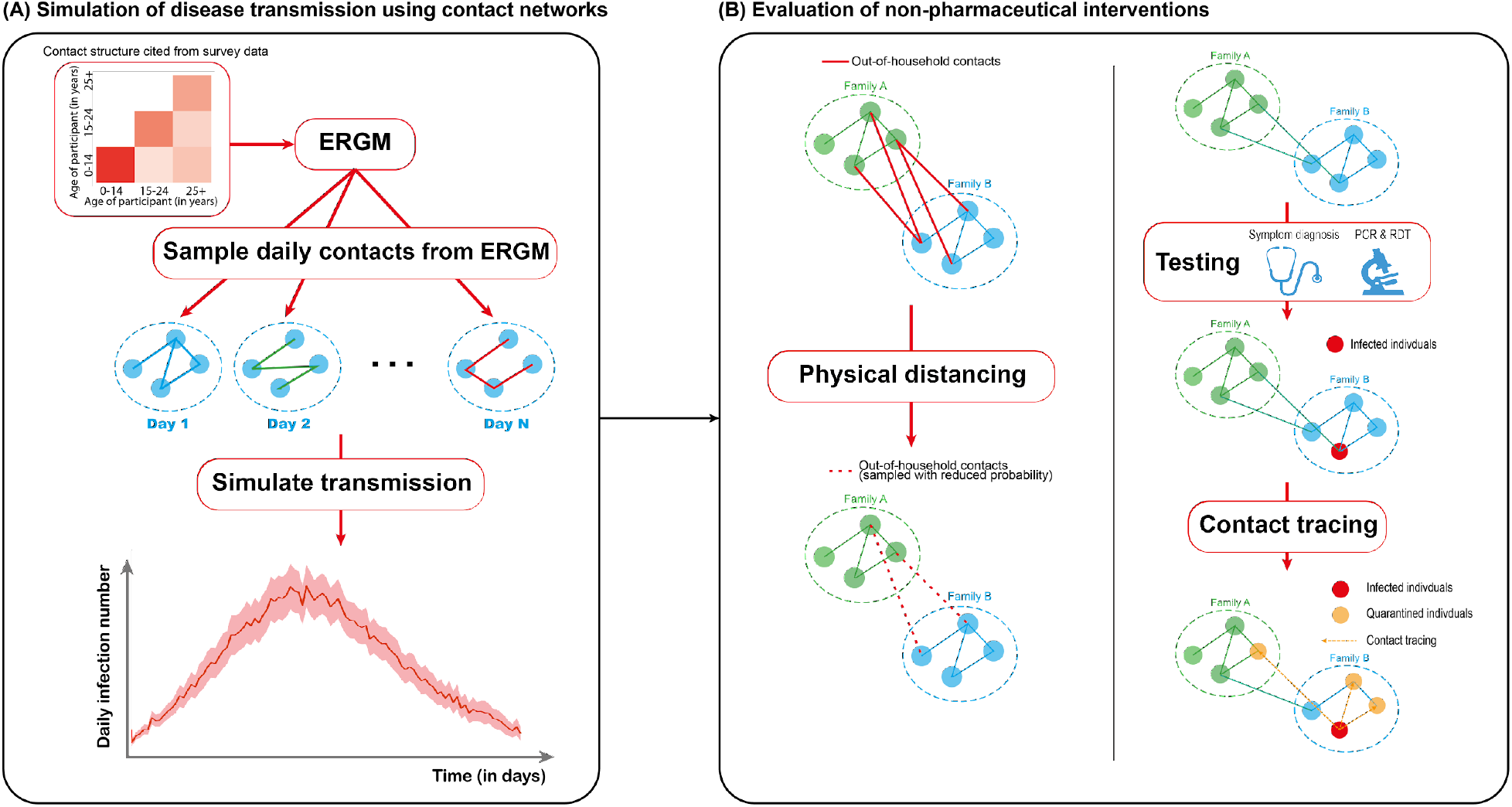
Schematic representation of methodology. A) For each parameter setting we simulated 20 synthetic populations and inferred the contact network with ERGM, using age- and household-structured contact data. At each simulation step, a contact network is sampled from the ERGM, where we simulate the transmission of SARS-CoV-2. B) We evaluated the effect of physical distancing by reducing the sampling probability of non-household contacts. Testing, isolation, contact tracing, and quarantine are also simulated using the contact networks.

We tested if our model reflects the cited contact data and transmission dynamics by comparing the simulated contact distribution and generation time with the empirical data. (Figure 2) The age mixing contact matrix generated from ERGM is also compared to the survey data. (Supplementary Figure 1) We performed simulations that covered the range of contact numbers collected from Africa CDC. (Figure 3) We cautioned these contact numbers are sampled from a small proportion of the populations and should not be considered as representing the contact number for each entire nation.

**Figure 2:**
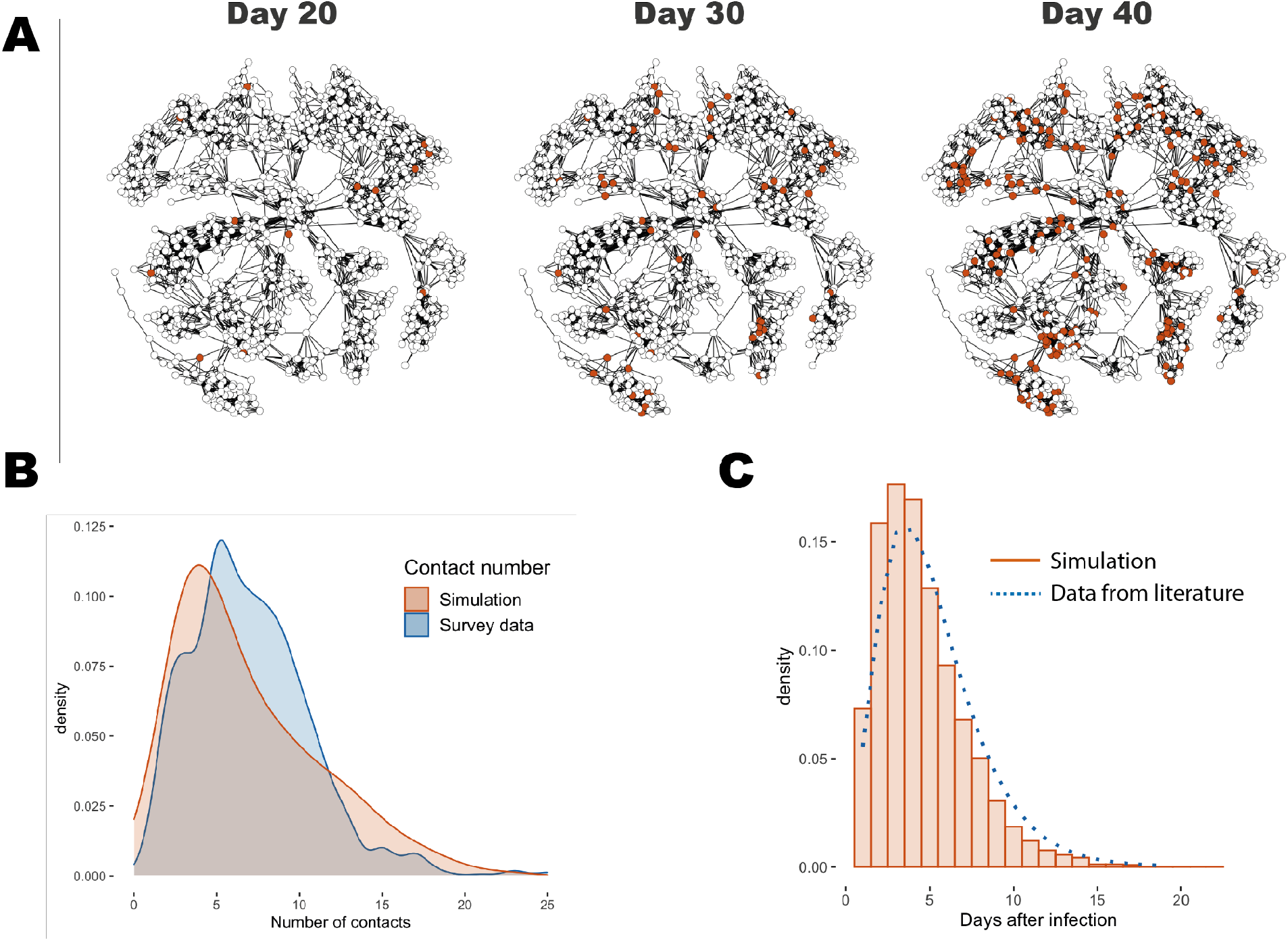
Properties of simulated outbreaks from the agent-based model. A) Example outbreak in a network composed of 1000 individuals, simulated under the urban setting without NPI. The left, middle and right panel show the distributions of infected individuals (red dots) on day 20, day 30 and day 40 from the day that initial cases are imported. For visualisation purposes we fix the network structure across days here, while in our analysis a different contact network is sampled each day. B) Comparison of the simulated contact number distribution with those computed from the cited Uganda survey data.^24^ The simulated contact number distribution is computed by sampling 100 daily contact realisations from each of the 20 synthetic populations. C) Comparison of generation time from simulation (red histogram) and that from cited Uganda survey data ^24^ (blue dotted curve).

**Figure 3:**
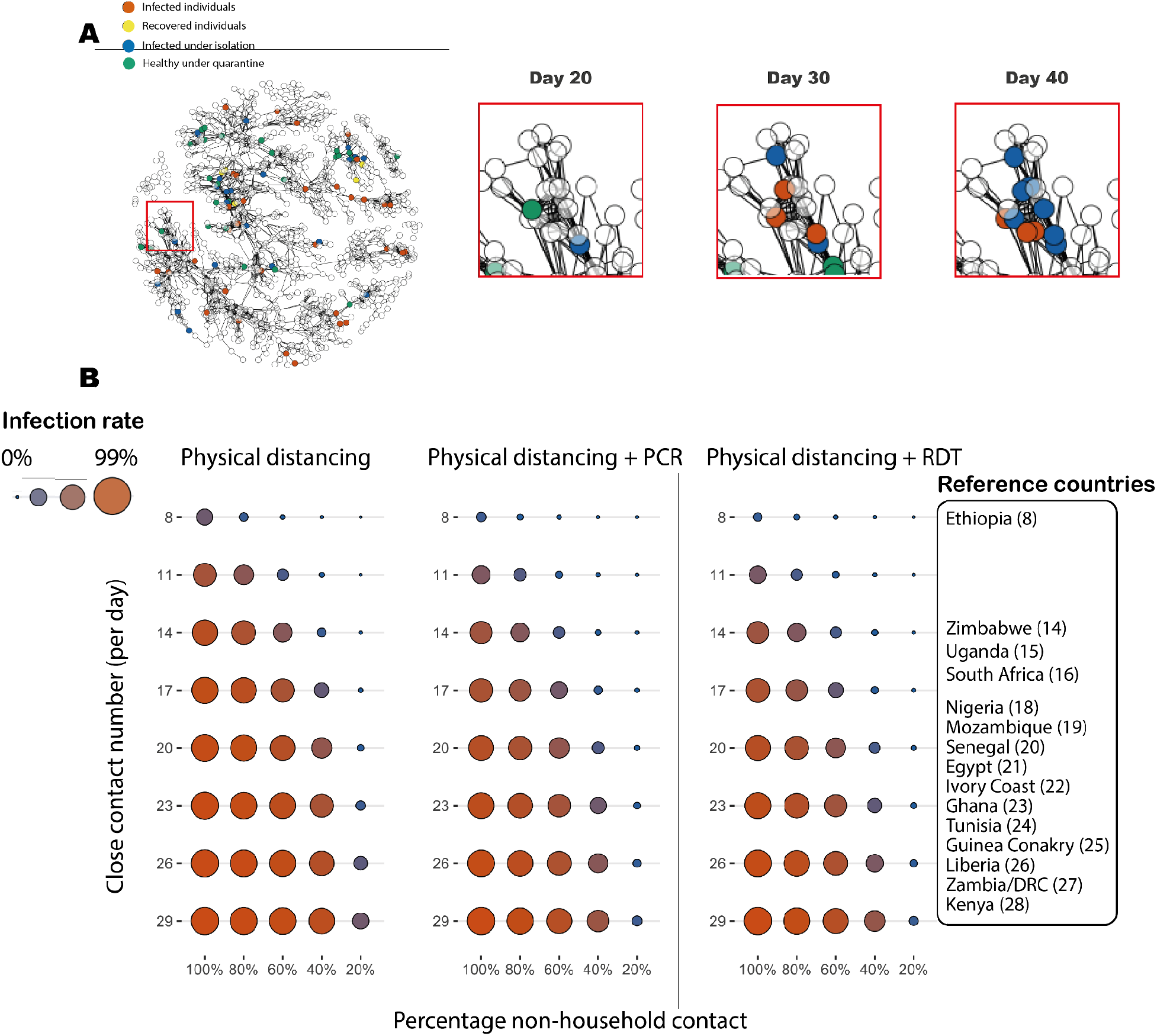
Evaluation of containing strategies. A) Example of case finding and quarantine in one simulated outbreak trajectory with NPIs, where a moderate physical distancing reduces 40% of the contact outside the household and symptom+PCR testing is performed. The network structure is fixed for visualisation, while in simulation a contact network is different each day. B) Social distancing and tests have different containing effects over different geographies. Synthetic populations were constructed using a grid of contact numbers that covers several reference LMICs. Left panel shows simulation with different levels of physical distance, indicated by the percentage of non-household contacts that are still permitted (no physical distancing: 100%; strongest physical distancing: 20%). The middle and right panel shows physical distancing combined with PCR or RDT tests respectively. The size and colour of the circle shows the mean of infection rate of 200 simulated outbreaks. Numbers and quantiles of the distribution are in Supplementary Table 2.

### NPI simulation

We evaluated the impact of testing alone and in combination with other NPIs, which include contact tracing, quarantine, and physical distancing, for LMICs. (Figure 1B) The testing methods considered were PCR and antigen RDT. We simulated that only individuals with symptoms and agree to be tested will receive testing. In our simulation, an infected individual with symptoms will be discovered at a rate which is the product of four parameters: healthcare seeking rate, test consent rate, sampling success rate and lab sensitivity of the test. Values and sources of testing parameters were summarised in Table 2.

**Table 2:** Summary of testing and NPI parameters. Last column shows the sources for cited parameters; values of assumed parameters are used in the containment simulations in Figure 3 and Figure 4, while separate sensitivity analyses are performed for each assumed parameter (Supplementary Table 3 and Supplementary Table 4). For rationales behind the chosen value for assumed parameters, see Supplementary Methods.

| Parameter | Symptom | PCR | Antigen RDT | Data source |
| --- | --- | --- | --- | --- |
| Test consent rate | 100% | 80% |  | Assumption |
| Isolation consent rate | 20% | 90% |  | Assumption |
| Days from symptom onset to isolation (days) | 1 | 5 | 1 | Assumption |
| Duration that close contacts are traced (days) | 3 | 5 | 3 | Assumption |
| Tracing success rate (non-household contacts) | 85% | 80% | 85% | Assumption |
| Quarantine consent rate of contacts | 50% | 70% |  | Assumption |
| Sampling success rate | NA | 70% |  | 29,42,43 |
| Lab sensitivity | NA | 100% | 80% (10%-90%) | 28,29,44 |
| Transmission rate after quarantine | Rural: 10%; Urban: 10%; High density: 20% |  |  | Assumption |

Our simulation assumes a test is always performed with close contact tracing and quarantine, unless specified otherwise. We assumed only close contacts of confirmed cases who consent to be isolated will be traced. Therefore, the overall quarantine rate is determined by the product of three parameters: isolation consent rate of the infector, tracing success rate (we assumed a tracing success rate equal to 100% for members within the same household of the infector), and quarantine consent rate of the contacts. Physical distancing was modelled by reducing the number of contacts outside the households. We simulated the effect of physical distancing by reducing the non-household contact number by 20% to 80% with 20% increments. We approximate the effective reproduction number Re under physical distancing by scaling Re proportionally to the average daily contact numbers.

In addition to individual containment measures, we also evaluated combined strategies based on current practice of COVID-19 testing. The combined strategies evaluated included 1) symptom-based isolation + PCR testing, 2) symptom-based isolation + antigen RDT, 3) and symptom-based isolation + PCR testing + antigen RDT. (Supplementary Table 3) In practice, it does not introduce extra cost to suggest individuals who show up at the test centre with symptoms to self isolate. Therefore we assumed all three containment strategies are accompanied by symptom based isolation, which has a low compliance rate (20%). All three strategies were replicated with and without physical distancing for 200 outbreak trajectories to estimate the average daily incidence number. We simulated each trajectory for 100 days to guarantee that the transmission extincts within the synthetic population.

### Sensitivity analysis

We performed sensitivity analyses for a list of parameters that could vary (Supplementary Table 4) and used regression to evaluate the impact of changing these parameters on the epidemic sizes. For the baseline conditions, we simulated outbreaks over grids of values for transmission and demographic variables that are assumed or could not be determined from literature. We computed R^2^ using the total infected number as the dependent variable to capture the proportion of variance explained by each variable. (Table 1). For the intervening conditions, we performed the same simulations over grids of values for testing and NPI variables. We computed the R^2^ using infected numbers under three different intervention strategies as the dependent variables and testing/NPI variables in Table 2 as independent variables. In total we have 2,028 simulation settings (3 community settings, 4 containing strategies and 169 values for the variables) which are listed in Supplementary table 5. For each simulation setting, we simulated 200 simulated trajectories of 100 days to obtain the bootstrap confidence interval (BCI) of the estimated infection numbers.

The code generated during this study is bundled up, with an R-package available at: https://github.com/Xilin-Jiang/NetworkCOVID19. Please refer to the GitHub page for installing the package and setting parameters for replicating our results or performing other contact network based analysis.

## Results

### Effectiveness of testing and physical distancing

To investigate the effectiveness of testing and physical distancing under different contact numbers based on survey data from African CDC. ^21^ We found moderate physical distancing (permitting 60% of non-household contacts) could contain the outbreaks for geographies when the contact number is below 11, with infection rate equal to 15.4% [95% BCI 13.5% - 17.2%]. (Figure 3B) Notably, for majority of LMIC countries considered, which have more than 20 close contacts per day, even strong physical distancing alone (permitting only 40% of non-household contacts) could not protect more than half of the population (infection rate = 53.5% [95% BCI 50.9% - 56.0%]). Only strict lockdown that reduces non-household contact number to 20% of that of normal could contain the outbreaks. (contact number = 20: infection rate = 5.1% [95% BCI 4.4% - 5.9%])

RDT or PCR alone could only mitigate the transmission in communities with less than 20 contacts, while combined with physical distancing, the mitigation effect increases, with 34.8% [95% BCI 33.3% - 36.3%] infection rate under moderate physical distancing. (contact number = 20, non-household contact rate= 60%) The mitigating effect increases as physical distancing gets stronger, with RDT slightly outperforming PCR tests. When simulating with a contact rate of 13, we found that using PCR or antigen RDT alone will isolate or quarantine more than 25% of the population and reduce the proportion of population infected as the isolation compliance rate increases. (Supplementary Figure 2A) When isolation compliance rates are at optimistic levels (90%), antigen RDT testing outperformed PCR and reduced the infection rate to 47.4% [95% BCI 45.8% - 49.1%].

### Combined strategies have varying performance in rural and urban communities

Motivated by the improved containment effect of combining tests with social distancing, we further evaluated combined strategies based on practice. Firstly, we assume patients who show up at the test centre with symptoms will be advised to isolate and told their contact to isolate (symptome-based isolation), which is modelled at a low compliance rate (Table 2); Secondly, we assumed a patient could provide sample for both a RDT and a PCR test. To contrast the contact structure between populations, we focused on three representative communities abstracted from contact data. We found that the symptom+PCR has approximately similar performance as symptom+RDT, while symptom+PCR+RDT has better performance in all three community settings. (Figure 4A, Supplementary Figure 3) In rural settings, either symptom+PCR or symptom+RDT could contain the outbreaks (symptom+PCR: 4.3% infected [95% BCI: 3.6% - 4.9%], Supplementary Table 3). In urban settings, symptom+PCR+RDT could suppress infection rate to 32.6% [95% BCI: 31.4% - 33.7%], while with physical distancing reduces non-household contact number by 40%, either symptom+PCR or RDT could contain the outbreaks. Notably, physical distancing could effectively flatten the curve in urban communities, (Supplementary Figure 3D) while the effect is less prominent in high density communities. Combined, these results show that affordable options such as RDT could be sufficient for low-density geographies similar to the rural settings, whereas testing and tracing programmes need to be combined with physical distancing to achieve containment in more densely populated areas similar to the urban settings.

**Figure 4:**
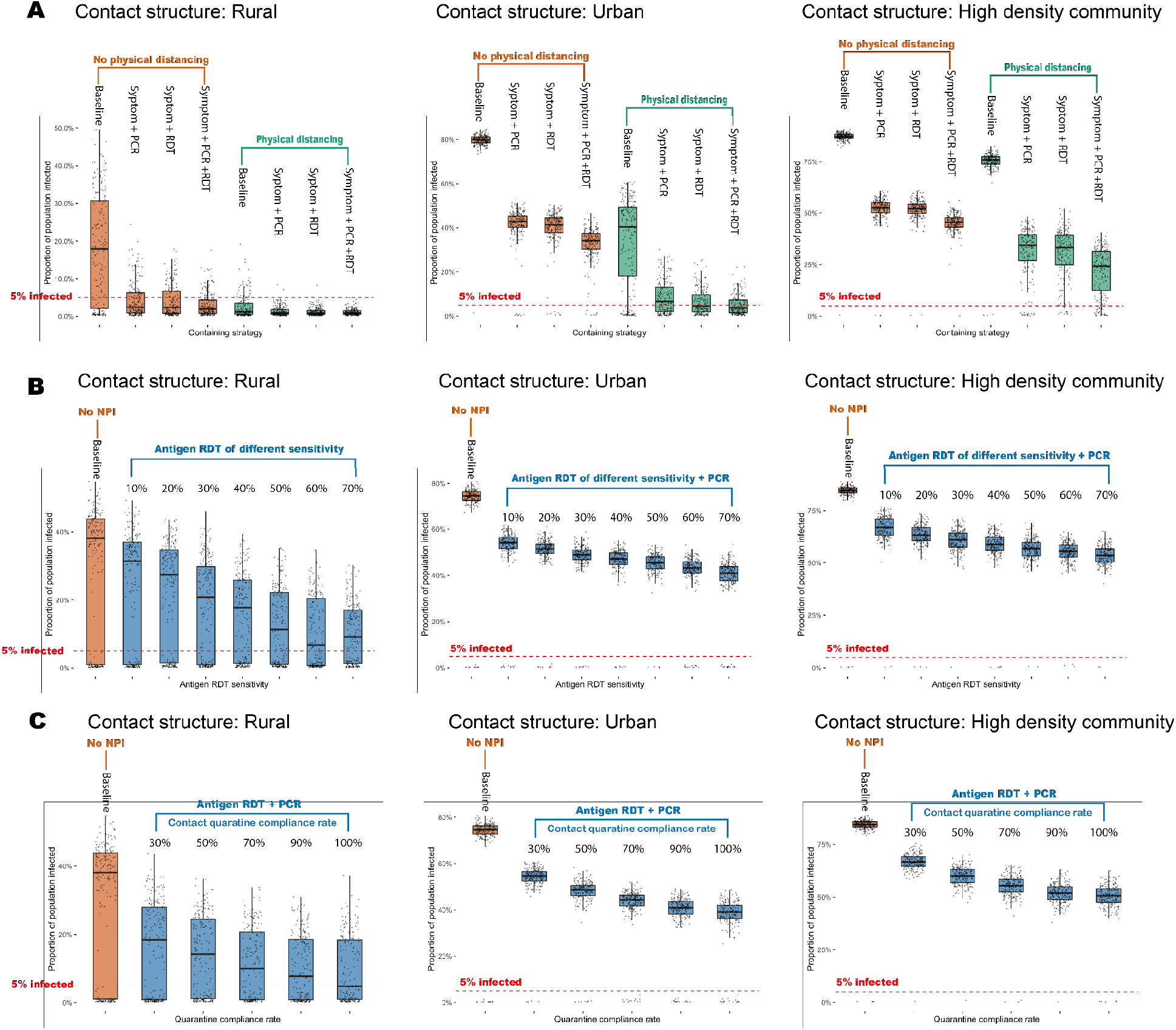
The impacts of testing strategies, sensitivity, and compliance rate on epidemic size vary with community settings. A) Infection rates under different combined strategies and in three abstracted community settings. Each dot represents one 100-day simulated outbreak and each box presents 200 simulated outbreaks, throughout all panels. Green boxes represent simulation with physical distancing (60% non-household contact permitted) and orange boxes show those without physical distancing. The quintiles of infection rate and mortality rate are summarised in Supplementary Table 3. B) Infection rates for each community setting when applying antigen RDT of various sensitivity. The blue boxes show the infection rate when applying the most effective testing strategy identified in the upper panel, with different antigen RDT sensitivity. C) Infection rates for each community setting when different quarantine compliances rates of traced contacts are simulated. The blue boxes show the infection rates when simulating containment using the most effective testing strategy identified in the first panel. In all three panels, one dot represents the epidemic size of one bootstrap trajectory. The red dashed line shows an infection rate at 5%, dots below which are defined as stochastic extinction events.

The distributions of infection rates show substantial uncertainty for rural communities and for urban communities when physical distancing is in place. (Figure 4A, Supplementary Table 3) These uncertainties suggest the outbreaks might have substantial variation in the epidemic sizes even for communities with similar demographics. To quantify the probability that imported cases do not start an outbreak, we defined the stochastic extinction events as the trajectories that infected less than 5% of the population. Under the three settings considered, the probability of stochastic extinction increases when testing, isolation and quarantine are implemented. In rural settings, symptom+RDT would increase the probability of stochastic extinction from 29% to 70%. In urban settings, the stochastic extinction events happened in 62% of the trajectories when physical distancing and symptom+RDT+PCR are implemented, which is a much larger proportion compared to 0.5% when no NPI is implemented. In the high density settings, the stochastic extinction probability remains low even when physical distancing and symptom+RDT+PCR are all implemented (13.5%). We verified that the probability of stochastic extinction does not depend on simulated population size. (Supplementary figure 4) The possibility of achieving high stochastic extinction rates in rural and urban settings suggest consistent testing, tracing, and physical distancing could reduce the probability of full scale outbreaks resulting from imported cases.

### The impact of RDT sensitivity, tracing and assumed parameters

Sampling success rate, antigen loading, and kit technology could all impact on the sensitivity of antigen RDT. ^28,29^ When simulating combined strategies with varying antigen RDT sensitivity, our simulations show that the containment effect increases as the RDT sensitivity increases. (Figure 4B) To reach a similar containment effect as in Figure 4A, which assumes an antigen RDT sensitivity of 56% (70% sampling success rate and 80% kit sensitivity), the kits deployed should have a sensitivity above 40%.

For the role of contact tracing, we found when the quarantine compliance rate increased from 30% to 100% under the symptom+RDT+PCR strategy, the average infection rate reduced from 15.7% to 9.5% in rural settings, from 53.2% to 36.5% in urban settings, and from 66.5% to 49.5% in high density settings. (Figure 4C) Note only contacts that are of isolated infectors that are successfully traced are subject to the quarantine. Without close contact quarantine measures, the combination strategies show no significant effect on reducing infection numbers (Supplementary Figure 5). Moreover, we found that the number of infected individuals discovered by testing is less than those discovered by contact tracing when the contact quarantine compliance rate is above 25%. (Supplementary Figure 6A)

The impact of other testing parameters on the containment outcomes are summarised in Supplementary table 3 using R^2^ and P-values (Supplementary Methods Section 4). Among these parameters, strong correlation between isolation consent rates and containing effectiveness are observed for all strategies and settings. Days from symptom onset to isolation, quarantine consent rate of contacts and tracing success rate are correlated with containing effectiveness in urban and high density settings, while less impactful in the rural settings. Additionally, we found that starting testing and quarantine after 10% of the population are infected could protect 31% of the healthy population under urban settings and 21% of the healthy population under the high density settings (Supplementary Figure 4B).

## Discussion

After two years since the beginning of the COVID-19 pandemic, special attention needs to be paid to LMICs as they suffer the most from global health inequality. Low vaccination rates and vulnerable health care systems in these countries could lead to overwhelming outbreaks and harbour new variants. ^6,7,30^ Our primary goal is to prioritise tools for LMICs to protect themselves against COVID-19, where we highlighted four interconnected elements: community knowledge, RDT, physical distancing, and contact tracing. These elements could be utilised to address two pressing challenges in LMICs. Firstly, how to design a cost-effective strategy for LMICs to reduce the financial burden of controlling the transmission of COVID-19? Secondly, how to design setting-specific surveillance or containment programmes that account for geographical variability in LMICs ^17,31,32^? We argue that community knowledge about demographics and contact structure is a valuable resource that could facilitate government decision making. For example, if a community is known to be densely populated and have crowded households, our results suggest that physical distancing is expected to be less effective there. As another example, if survey data suggested some communities have less indoor contacts than average, we would expect a light containment strategy such as mild physical distancing alone could work effectively, thus saving resources from an over-stringent strategy. In practice, policy makers could perform contact surveys in representative communities and use them to construct a classification system. Our analyses present the trends of transmission dynamics and containment effectiveness across different contact numbers and demographics. When used in combination with community knowledge, our results could guide policy design that achieves the best outcomes while saving resources.

Antigen RDT has several desirable properties for application in LMICs. It is cheap, easy to distribute, has quick turnarounds, and doesn’t require lab facilities. LMICs require consistent containment to protect their health systems from being overwhelmed, which makes RDT an ideal choice to avoid huge financial costs. Specifically, RDT might be prioritised in resource-limited regions where contact number is low and performing PCR tests is prohibitive. Though containing power subjects to the test sensitivity and community scenario, our results suggested high quality kits that have a sensitivity above 50% will provide similar containment power shown in the simulation. However, the detrimental effect of false negative and false positive results might negate the containment effects. ^33^ We suggest real world RDT application should be accompanied by careful sensitivity and false positivity evaluation.

We also want to emphasise that policy design should serve its goals. If aiming at a zero-COVID policy, such as those deployed by China (as of March 5th, 2022), our results show that contact tracing is an essential element to implement, which relies on abundant personnel and quarantine capacities. However, if the goal is to flatten the curve to avoid overwhelmed health systems, mild-physical distancing is an effective measure which saves resources and might serve the young population of LMICs better. If the goal is to monitor the transmission for early alarm of new variants and prepare for outbreaks, mass RDT tests could detect abnormal outbreaks that link to new variants and auxiliary PCR tests could monitor prevalence of each variant. Successful containment strategy should apply community knowledge to serve the goal, whether it is suppressing, delaying, or monitoring the transmission of COVID-19.

There are some limitations in our study. Firstly, our simulated population is an approximation to the communities in LMICs and is limited by the available empirical evidence from LMICs.

Therefore, to apply our results to specific geography requires knowledge of the target population, including demographic and contact information. Secondly, like many simulation studies, we had to choose our parameters from empirical studies that are not consistent with each other. ^34^ This difficulty is most pronounced when we are setting the parameters for age-dependent susceptibility and asymptomatic rate of the infected. Lastly, Compared to simulations on larger networks that involve more sophisticated configuration such as schools and shopping malls, ^35^ our strategy might not capture superspreading events that were reported. Future development of approximation methods might provide comparable accuracy while scaling to large populations.

## Contributors

All authors contributed to study design. XJ designed the model, XJ and WG implemented the model, and XJ, WG, CF and CH analysed and verified data. All authors interpreted results, XJ, CF and CH drafted the manuscript, XJ made the figures, and all authors critically revised and approved the manuscript.

## Code availability

The code generated during this study is available at: https://github.com/Xilin-Jiang/NetworkCOVID19

## Acknowledgement

Computation used the Oxford Biomedical Research Computing (BMRC) facility, a joint development between the Wellcome Centre for Human Genetics and the Big Data Institute supported by Health Data Research UK and the NIHR Oxford Biomedical Research Centre. The views expressed are those of the authors and not necessarily those of the NHS, the NIHR or the Department of Health. We thank Microsoft China Co. Ltd. for computation power during the exploration stage of the study. We thank Yang Liu, Jiayao Lei and Mark Jit for discussion and comments on the manuscript.

## Supplementary Tables and Figures

**Supplementary Table 1:** Summary of the demographic information used in the study. We used the age and household size distributions from Uganda, except for the household size distribution in high density communities, where we used data from Afghanistan, which has the largest average household size recorded by the United Nations. ^18,19^

| Age group (in years) | Uganda |  |
| --- | --- | --- |
| 0-14 | 48.10% |  |
| 15-24 | 20.30% |  |
| 25+ | 31.60% |  |
| Household Size (members) | Uganda | High density community |
| 1 | 11.00% | 0.00% |
| 2-3 | 22.00% | 6.00% |
| 4-5 | 27.00% | 17.00% |
| 6+ | 40.00% | 77.00% |

**Supplementary Table 2:** Summary of the distributions of infection rates for that of Figure 3B. The mean of infection rates is computed from 200 simulated trajectories of 100 days for each setting. The range in the bracket shows the 10% and 90% quantiles of the infection rates from the 200 bootstrap sample trajectories.

| Contact number | Physical distancing |  |  |  |  | Physical distancing + PCR |  |  |  |  | Physical distancing + RDT |  |  |  |  |
| --- | --- | --- | --- | --- | --- | --- | --- | --- | --- | --- | --- | --- | --- | --- | --- |
|  | 100% | 80% | 60% | 40% | 20% | 100% | 80% | 60% | 40% | 20% | 100% | 80% | 60% | 40% | 20% |
| 8 | 31.8%<br>(0.5% - 51.1%) | 9.0% (0.3% - 25.9%) | 2.0% (0.3% - 5.1%) | 0.8% (0.2% - 1.6%) | 0.4% (0.2% - 0.8%) | 8.1% (0.5% - 17.4%) | 2.9% (0.3% - 7.6%) | 1.4% (0.3% - 3.0%) | 0.6% (0.2% - 1.3%) | 0.5% (0.2% - 0.9%) | 5.9% (0.4% - 15.2%) | 2.8% (0.4% - 7.3%) | 1.2% (0.3% - 2.6%) | 0.7% (0.2% - 1.4%) | 0.4% (0.2% - 0.7%) |
| 11 | 66.7%<br>(66.6% - 75.0%) | 48.1%<br>(1.3% - 62.2%) | 15.4%<br>(0.5% - 34.2%) | 2.5% (0.3% - 7.2%) | 0.7% (0.2% - 1.4%) | 29.9%<br>(13.3% - 39.0%) | 14.0%<br>(1.0% - 26.6%) | 4.4% (0.6% - 10.2%) | 1.4% (0.3% - 3.1%) | 0.6% (0.2% - 1.1%) | 26.2%<br>(7.7% - 37.7%) | 10.7%<br>(0.6% - 23.1%) | 3.5% (0.4% - 8.0%) | 1.1% (0.3% - 2.6%) | 0.6% (0.2% - 1.1%) |
| 14 | 82.5%<br>(80.2% - 85.7%) | 72.4%<br>(70.1% - 78.3%) | 45.5%<br>(0.5% - 62.2%) | 9.1% (0.4% - 21.4%) | 1.2% (0.3% - 2.8%) | 46.5%<br>(41.7% - 51.6%) | 32.5%<br>(20.1% - 43.4%) | 12.0%<br>(0.8% - 25.2%) | 2.7% (0.3% - 5.9%) | 0.9% (0.2% - 2.0%) | 43.4%<br>(37.9% - 50.4%) | 29.4%<br>(5.1% - 41.3%) | 10.4%<br>(1.0% - 24.0%) | 2.7% (0.4% - 6.1%) | 0.8% (0.2% - 1.6%) |
| 17 | 88.5%<br>(87.3% - 90.9%) | 83.5%<br>(81.1% - 85.8%) | 69.4%<br>(66.4% - 75.8%) | 26.5%<br>(1.0% - 47.9%) | 2.3% (0.3% - 5.8%) | 52.2%<br>(47.3% - 56.9%) | 44.6%<br>(39.0% - 51.0%) | 25.7%<br>(6.1% - 38.5%) | 6.1% (0.7% - 14.0%) | 1.3% (0.3% - 2.8%) | 51.2%<br>(46.0% - 55.7%) | 40.6%<br>(34.1% - 47.7%) | 19.4%<br>(1.9% - 34.1%) | 3.9% (0.4% - 8.4%) | 1.1% (0.3% - 2.3%) |
| 20 | 92.8%<br>(91.3% - 94.2%) | 88.8%<br>(86.8% - 90.6%) | 79.6%<br>(78.0% - 83.8%) | 53.5%<br>(27.4% - 67.4%) | 5.1% (0.3% - 13.2%) | 58.7%<br>(54.3% - 63.2%) | 52.2%<br>(47.6% - 56.6%) | 39.3%<br>(29.5% - 47.1%) | 13.0%<br>(1.5% - 25.7%) | 1.9% (0.3% - 4.5%) | 57.0%<br>(52.0% - 61.8%) | 49.9%<br>(45.5% - 55.0%) | 34.8%<br>(22.0% - 44.7%) | 10.1%<br>(1.0% - 20.5%) | 1.9% (0.4% - 4.2%) |
| 23 | 95.1%<br>(94.1% - 96.2%) | 92.1%<br>(90.7% - 93.5%) | 86.2%<br>(83.8% - 88.5%) | 69.4%<br>(65.3% - 76.8%) | 10.5%<br>(0.4% - 25.8%) | 63.5%<br>(60.1% - 67.3%) | 57.8%<br>(53.1% - 61.6%) | 48.8%<br>(43.6% - 53.4%) | 24.1%<br>(6.4% - 38.6%) | 3.4% (0.5% - 7.9%) | 61.6%<br>(56.7% - 65.7%) | 55.1%<br>(50.8% - 60.0%) | 44.6%<br>(37.1% - 51.7%) | 17.7%<br>(1.3% - 31.0%) | 2.7% (0.5% - 6.8%) |
| 26 | 96.6%<br>(95.6% - 97.5%) | 94.2%<br>(93.1% - 95.3%) | 89.0%<br>(88.0% - 91.7%) | 78.1%<br>(76.1% - 82.7%) | 22.7%<br>(0.7% - 44.3%) | 68.1%<br>(64.8% - 71.7%) | 62.3%<br>(58.9% - 65.9%) | 54.3%<br>(50.4% - 59.1%) | 34.5%<br>(15.8% - 46.2%) | 5.0% (0.5% - 11.3%) | 65.2%<br>(61.1% - 68.8%) | 60.0%<br>(56.3% - 63.6%) | 51.6%<br>(46.4% - 56.9%) | 27.2%<br>(7.5% - 41.2%) | 4.3% (0.5% - 9.9%) |
| 29 | 97.6%<br>(96.9% - 98.3%) | 95.8%<br>(94.9% - 96.9%) | 92.3%<br>(91.0% - 93.8%) | 85.0%<br>(82.6% - 87.1%) | 33.1%<br>(1.1% - 57.9%) | 72.0%<br>(69.0% - 74.9%) | 66.2%<br>(62.3% - 69.8%) | 59.5%<br>(55.4% - 63.5%) | 44.4%<br>(36.3% - 52.4%) | 8.0% (1.0% - 17.3%) | 68.6%<br>(64.8% - 72.2%) | 63.5%<br>(59.3% - 67.9%) | 56.8%<br>(51.5% - 61.2%) | 38.9%<br>(22.1% - 48.8%) | 6.8% (0.8% - 13.7%) |

**Supplementary Table 3:** Summary of the distributions of infection and mortality rate for three community settings. The mean of infection rate and mortality rates is computed from 200 simulated trajectories of 100 days for each strategy. The range in the bracket shows the 10% and 90% quantiles of the infection rates from 200 bootstrap samples.

| Strategies | Infection rate |  |  | Mortality rate |  |  |
| --- | --- | --- | --- | --- | --- | --- |
|  | Rural | Urban | High density | Rural | Urban | High density |
| No intervention | 18.0% (0.5% - 38.2%) | 79.5% (77.3% - 82.4%) | 87.0% (85.2% - 89.4%) | 0.1% (0.0% - 0.2%) | 0.4% (0.2% - 0.6%) | 0.4% (0.1% - 0.7%) |
| Symptom + PCR | 4.3% (0.4% - 11.5%) | 41.0% (36.0% - 47.5%) | 51.8% (47.1% - 56.9%) | 0.0% (0.0% - 0.1%) | 0.2% (0.0% - 0.4%) | 0.3% (0.1% - 0.5%) |
| Symptom + RDT | 4.0% (0.4% - 10.3%) | 39.7% (34.1% - 46.9%) | 51.6% (47.5% - 56.8%) | 0.0% (0.0% - 0.1%) | 0.2% (0.0% - 0.4%) | 0.3% (0.1% - 0.5%) |
| Symptom + PCR + RDT | 3.0% (0.4% - 7.6%) | 32.6% (25.4% - 39.8%) | 45.3% (40.9% - 50.3%) | 0.0% (0.0% - 0.1%) | 0.2% (0.0% - 0.4%) | 0.2% (0.0% - 0.4%) |
| Reducing 40% of non-household contact | 2.5% (0.3% - 6.9%) | 33.7% (0.8% - 55.5%) | 74.1% (71.6% - 78.9%) | 0.0% (0.0% - 0.1%) | 0.2% (0.0% - 0.4%) | 0.4% (0.2% - 0.6%) |
| Symptom + PCR + reducing 40% of non-household contact | 1.3% (0.3% - 3.2%) | 8.4% (0.7% - 19.7%) | 31.1% (7.0% - 43.0%) | 0.0% (0.0% - 0.0%) | 0.0% (0.0% - 0.1%) | 0.1% (0.0% - 0.3%) |
| Symptom + RDT + reducing 40% of non-household contact | 1.2% (0.3% - 2.7%) | 6.5% (0.6% - 14.4%) | 29.8% (6.1% - 42.4%) | 0.0% (0.0% - 0.0%) | 0.0% (0.0% - 0.1%) | 0.1% (0.0% - 0.3%) |
| Symptom + PCR + RDT + reducing 40% of non-household contact | 1.1% (0.3% - 2.2%) | 5.0% (0.6% - 11.8%) | 21.6% (3.2% - 34.4%) | 0.0% (0.0% - 0.0%) | 0.0% (0.0% - 0.1%) | 0.1% (0.0% - 0.2%) |

**Supplementary Table 4:**
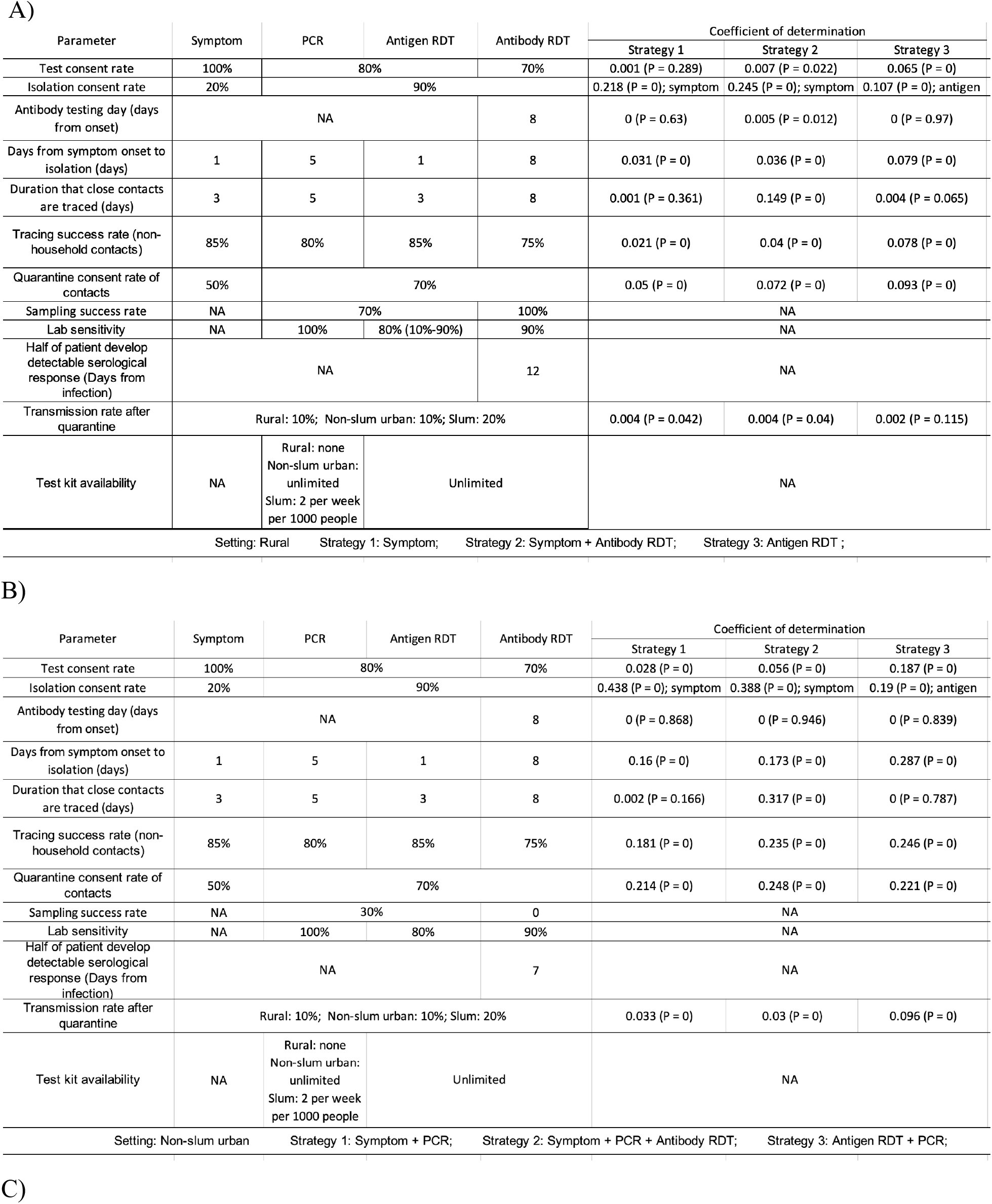
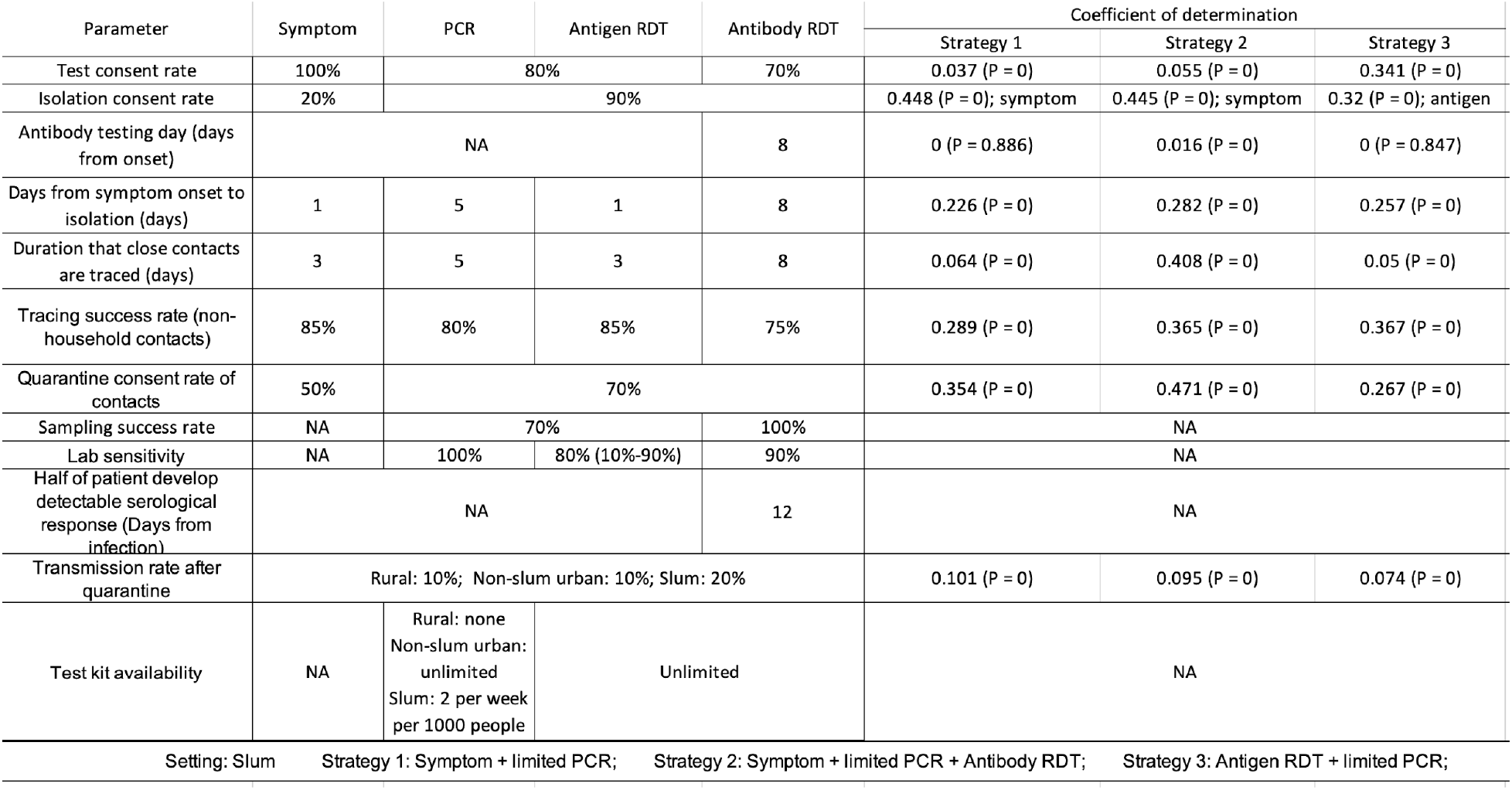
The impact of NPI parameter setting on the effectiveness of containment. Panel A-C shows sensitivity analysis under rural, urban and high density settings. The coefficient of determination (R^2^) shows the proportion of variance in the final infected number (under each containing strategy) that is captured by the testing parameters; The P-values in the bracket show the rate of type one error of rejecting the null hypothesis that the parameter is not correlated with outcome.

| Parameter | Symptom | PCR | Antigen RDT | Antibody RDT | Coefficient of determination |  |  |
| --- | --- | --- | --- | --- | --- | --- | --- |
|  |  |  |  |  | Strategy 1 | Strategy 2 | Strategy 3 |
| Test consent rate | 100% | 80% |  | 70% | 0.001 (P = 0.289) | 0.007 (P = 0.022) | 0.065 (P = 0) |
| Isolation consent rate | 20% | 90% |  |  | 0.218 (P = 0); symptom | 0.245 (P = 0); symptom | 0.107 (P = 0); antigen |
| Antibody testing day (days from onset) | NA |  |  | 8 | 0 (P = 0.63) | 0.005 (P = 0.012) | 0 (P = 0.97) |
| Days from symptom onset to isolation (days) | 1 | 5 | 1 | 8 | 0.031 (P = 0) | 0.036 (P = 0) | 0.079 (P = 0) |
| Duration that close contacts are traced (days) | 3 | 5 | 3 | 8 | 0.001 (P = 0.361) | 0.149 (P = 0) | 0.004 (P = 0.065) |
| Tracing success rate (non-household contacts) | 85% | 80% | 85% | 75% | 0.021 (P = 0) | 0.04 (P = 0) | 0.078 (P = 0) |
| Quarantine consent rate of contacts | 50% | 70% |  |  | 0.05 (P = 0) | 0.072 (P = 0) | 0.093 (P = 0) |
| Sampling success rate | NA | 70% |  | 100% | NA |  |  |
| Lab sensitivity | NA | 100% | 80% (10%-90%) | 90% | NA |  |  |
| Half of patient develop detectable serological response (Days from infection) | NA |  |  | 12 | NA |  |  |
| Transmission rate after quarantine | Rural: 10%; Non-slum urban: 10%; Slum: 20% |  |  |  | 0.004 (P = 0.042) | 0.004 (P = 0.04) | 0.002 (P = 0.115) |
| Test kit availability | NA | Rural: none<br>Non-slum urban: unlimited<br>Slum: 2 per week per 1000 people | Unlimited |  | NA |  |  |
| Setting: Rural Strategy 1: Symptom; Strategy 2: Symptom + Antibody RDT; Strategy 3: Antigen RDT ; |  |  |  |  |  |  |  |

B)
| Parameter | Symptom | PCR | Antigen RDT | Antibody RDT | Coefficient of determination |  |  |
| --- | --- | --- | --- | --- | --- | --- | --- |
|  |  |  |  |  | Strategy 1 | Strategy 2 | Strategy 3 |
| Test consent rate | 100% | 80% |  | 70% | 0.028 (P = 0) | 0.056 (P = 0) | 0.187 (P = 0) |
| Isolation consent rate | 20% | 90% |  |  | 0.438 (P = 0); symptom | 0.388 (P = 0); symptom | 0.19 (P = 0); antigen |
| Antibody testing day (days from onset) | NA |  |  | 8 | 0 (P = 0.868) | 0 (P = 0.946) | 0 (P = 0.839) |
| Days from symptom onset to isolation (days) | 1 | 5 | 1 | 8 | 0.16 (P = 0) | 0.173 (P = 0) | 0.287 (P = 0) |
| Duration that close contacts are traced (days) | 3 | 5 | 3 | 8 | 0.002 (P = 0.166) | 0.317 (P = 0) | 0 (P = 0.787) |
| Tracing success rate (non-household contacts) | 85% | 80% | 85% | 75% | 0.181 (P = 0) | 0.235 (P = 0) | 0.246 (P = 0) |
| Quarantine consent rate of contacts | 50% | 70% |  |  | 0.214 (P = 0) | 0.248 (P = 0) | 0.221 (P = 0) |
| Sampling success rate | NA | 30% |  | 0 | NA |  |  |
| Lab sensitivity | NA | 100% | 80% | 90% | NA |  |  |
| Half of patient develop detectable serological response (Days from infection) | NA |  |  | 7 | NA |  |  |
| Transmission rate after quarantine | Rural: 10%; Non-slum urban: 10%; Slum: 20% |  |  |  | 0.033 (P = 0) | 0.03 (P = 0) | 0.096 (P = 0) |
| Test kit availability | NA | Rural: none<br>Non-slum urban: unlimited<br>Slum: 2 per week per 1000 people | Unlimited |  | NA |  |  |
| Setting: Non-slum urban Strategy 1: Symptom + PCR; Strategy 2: Symptom + PCR + Antibody RDT; Strategy 3: Antigen RDT + PCR; |  |  |  |  |  |  |  |
|  |  |  |  |  | Strategy 1 | Strategy 2 | Strategy 3 |
| Test consent rate | 100% | 80% |  | 70% | 0.037 (P = 0) | 0.055 (P = 0) | 0.341 (P = 0) |
| Isolation consent rate | 20% | 90% |  |  | 0.448 (P = 0); symptom | 0.445 (P = 0); symptom | 0.32 (P = 0); antigen |
| Antibody testing day (days from onset) | NA |  |  | 8 | 0 (P = 0.886) | 0.016 (P = 0) | 0 (P = 0.847) |
| Days from symptom onset to isolation (days) | 1 | 5 | 1 | 8 | 0.226 (P = 0) | 0.282 (P = 0) | 0.257 (P = 0) |
| Duration that close contacts are traced (days) | 3 | 5 | 3 | 8 | 0.064 (P = 0) | 0.408 (P = 0) | 0.05 (P = 0) |
| Tracing success rate (non-household contacts) | 85% | 80% | 85% | 75% | 0.289 (P = 0) | 0.365 (P = 0) | 0.367 (P = 0) |
| Quarantine consent rate of contacts | 50% | 70% |  |  | 0.354 (P = 0) | 0.471 (P = 0) | 0.267 (P = 0) |
| Sampling success rate | NA | 70% |  | 100% | NA |  |  |
| Lab sensitivity | NA | 100% | 80% (10%-90%) | 90% | NA |  |  |
| Half of patient develop detectable serological response (Days from infection) | NA |  |  | 12 | NA |  |  |
| Transmission rate after quarantine | Rural: 10%; Non-slum urban: 10%; Slum: 20% |  |  |  | 0.101 (P = 0) | 0.095 (P = 0) | 0.074 (P = 0) |
| Test kit availability | NA | Rural: none<br>Non-slum urban: unlimited<br>Slum: 2 per week per 1000 people | Unlimited |  | NA |  |  |
| Setting: Slum Strategy 1: Symptom + limited PCR; Strategy 2: Symptom + limited PCR + Antibody RDT; Strategy 3: Antigen RDT + limited PCR; |  |  |  |  |  |  |  |

Setting: Slum Strategy 1: Symptom + limited PCR; Strategy 2: Symptom + limited PCR + Antibody RDT; Strategy 3: Antigen RDT + limited PCR;

**Supplementary Table 5:** Parameter settings for sensitivity analysis. The Description explains what variables are changed and rows contain the values of each variable used for simulation. For each value we performed the simulation for baseline and three types of containing strategies under the rural, urban and high density settings.

| ID | Description | Values |  |  |  |  |  |  |  |  |
| --- | --- | --- | --- | --- | --- | --- | --- | --- | --- | --- |
| 1 | Basic reproduction number | 1.5 | 2 | 2.5 | 3 | 3.5 |  |  |  |  |
| 2 | Number of daily contact | 2 | 4 | 7 | 10 | 14 |  |  |  |  |
| 3 | Number of initial importing cases | 1 | 2 | 5 | 8 | 15 |  |  |  |  |
| 4 | Healthcare seeking rate | 30% | 50% | 70% | 90% | 100% |  |  |  |  |
| 5 | Asymptomatic rate | 10% | 20% | 30% | 40% | 50% | 60% | 70% | 80% | 90% |
| 6 | Transmission ratio for isolated patients | 5% | 10% | 15% | 20% | 25% |  |  |  |  |
| 7 | Daily PCR tests per 1000 population for slum | Not available | 1 | 2 | 10 | 20 | 50 | 1000 |  |  |
| 8 | Community size | 100 | 1000 | 2000 |  |  |  |  |  |  |
| 9 | Infection number in 1000 to trigger containment | No delay | 5 | 20 | 50 | 100 | 250 | 500 |  |  |
| 10 | Percentage of contacts that are with a member of household, fixing the number of total contacts | 20% | 40% | 60% (did not converge for slum settings) | 80% (did not converge for three settings) |  |  |  |  |  |
| 11 | Social distancing, which is computed as the proportion of non-household contact that is stopped. | 0% | 20% | 40% | 60% (did not converge for rural setting) | 80% (did not converge for rural and slum settings) |  |  |  |  |
| 12 | Non-household contact trace successful rate | 0% | 15% | 35% | 55% | 75% | 85% | 95% |  |  |
| 13 | Delay after tested positive to isolation (days) | 0 | 1 | 3 | 5 |  |  |  |  |  |
| 14 | Duration of contact tracing, counted from the day of isolation (days) | Symptom screening/Antigen RDT = 1; Antibody RDT = 6; PCR = 3 | Symptom screening/Antigen RDT = 3; Antibody RDT = 8; PCR = 5 | Symptom screening/Antigen RDT = 5; Antibody RDT = 10; PCR = 7 | Symptom screening/Antigen RDT = 7; Antibody RDT = 12; PCR = 9 |  |  |  |  |  |
| 15 | Delay after onset for PCR testing results (days); contact tracing duration is set to be same as the delay, as we assume healthcare workers will start contact tracing once they identify an individual with onset. | 0 | 1 | 3 | 5 | 7 | 10 | 14 |  |  |
| 16 | Delay after onset for antibody sampling (days); contact tracing duration is set to be same as the delay, as we assume healthcare workers will start contact tracing once they identify an individual with onset. | 5 | 6 | 7 | 8 | 9 | 10 | 11 |  |  |
| 17 | Seroconversion rate | optimistic: half of individual has detectable antibody 7 days after onset | optimistic: half of individual has detectable antibody 12 days after infection | conservative: half of individual has detectable antibody 9 days after onset |  |  |  |  |  |  |
| 18 | Test acceptance rate (conditional on an individual seek healthcare) | Antibody (blood sample) = 30%; PCR/Antigen (swab sample) = 40% | Antibody (blood sample) = 50%; PCR/Antigen (swab sample) = 60% | Antibody (blood sample) = 70%; PCR/Antigen (swab sample) = 80% | 90% |  |  |  |  |  |
| 19 | Symptom-based isolation consent rate, simulated with other testing measures | 10% | 20% | 30% | 40% | 50% | 60% | 70% | 80% | 90% |
| 20 | Symptom-based isolation consent rate (simulation with only symptom detection, no other detecting measures) | 10% | 20% | 30% | 40% | 50% | 60% | 70% | 80% | 90% |
| 21 | PCR isolation consent rate (simulation with only PCR testing, no other detecting measure) | 10% | 20% | 30% | 40% | 50% | 60% | 70% | 80% | 90% |
| 22 | Antibody RDT isolation consent rate (simulation with only antibody RDT, no other detecting measure) | 10% | 20% | 30% | 40% | 50% | 60% | 70% | 80% | 90% |
| 23 | Antigen RDT isolation consent rate (simulation with only antigen RDT, no other detecting measure) | 10% | 20% | 30% | 40% | 50% | 60% | 70% | 80% | 90% |
| 24 | close contact quarantine compliance rate (conditional on the infector consent to be isolated) | Symptom screen = 10%; Testing = 30% | Symptom screen = 30%; Testing = 50% | Symptom screen = 50%; Testing = 70% | Symptom screen = 70%; Testing = 90% | Symptom screen = 90%; Testing = 100% |  |  |  |  |

**Supplementary Table 6:** Lack-of-fit test for the ERGM inference. We performed lack-of-fit tests of the target statistics for the four variables that alter the ERGMs. IDs correspond to Supplementary Table 3. For each value (bold), we compute the P-value by comparing the 20 fitted networks with the input to the ERGMs. * shows the cases when the networks are rejected as good fits.

| ID | Description | Values |  |  |  |  |
| --- | --- | --- | --- | --- | --- | --- |
| 2 | Number of daily contact | 2 | 4 | 7 | 10 | 14 |
|  | Rural | 0.99 | 1.00 | 1.00 | 1.00 | 0.14 |
|  | Non-slum urban | 1.00 | 1.00 | 0.99 | 1.00 | 1.00 |
|  | Slum | 1.00 | 0.94 | 0.99 | 1.00 | 1.00 |
| 9 | Community size | 100 (did not converge for rural setting) | 1000 | 2000 |  |  |
|  | Rural | 0.000002 * | 1.00 | 1.00 |  |  |
|  | Non-slum urban | 1.00 | 1.00 | 1.00 |  |  |
|  | Slum | 1.00 | 1.00 | 1.00 |  |  |
| 11 | Percentage of contacts that are with a member of household, fixing the number of total contacts | 20% | 40% | 60% (did not converge for rural setting) | 80% (did not converge for three settings) |  |
|  | Rural | 1.00 | 1.00 | 1.00 | 0 * |  |
|  | Non-slum urban | 1.00 | 1.00 | 0.006* | 0 * |  |
|  | Slum | 0.99 | 1.00 | 1.00 | 0 * |  |
| 12 | Social distancing, which is computed as the proportion of non-household contact that is stopped. | 0% | 20% | 40% | 60% | 80% |
|  | Rural | 1.00 | 1.00 | 0.99 | 1.00 | 1.00 |
|  | Non-slum urban | 1.00 | 1.00 | 1.00 | 1.00 | 1.00 |
|  | Slum | 1.00 | 1.00 | 1.00 | 1.00 | 1.00 |

**Supplementary Figure 1:**
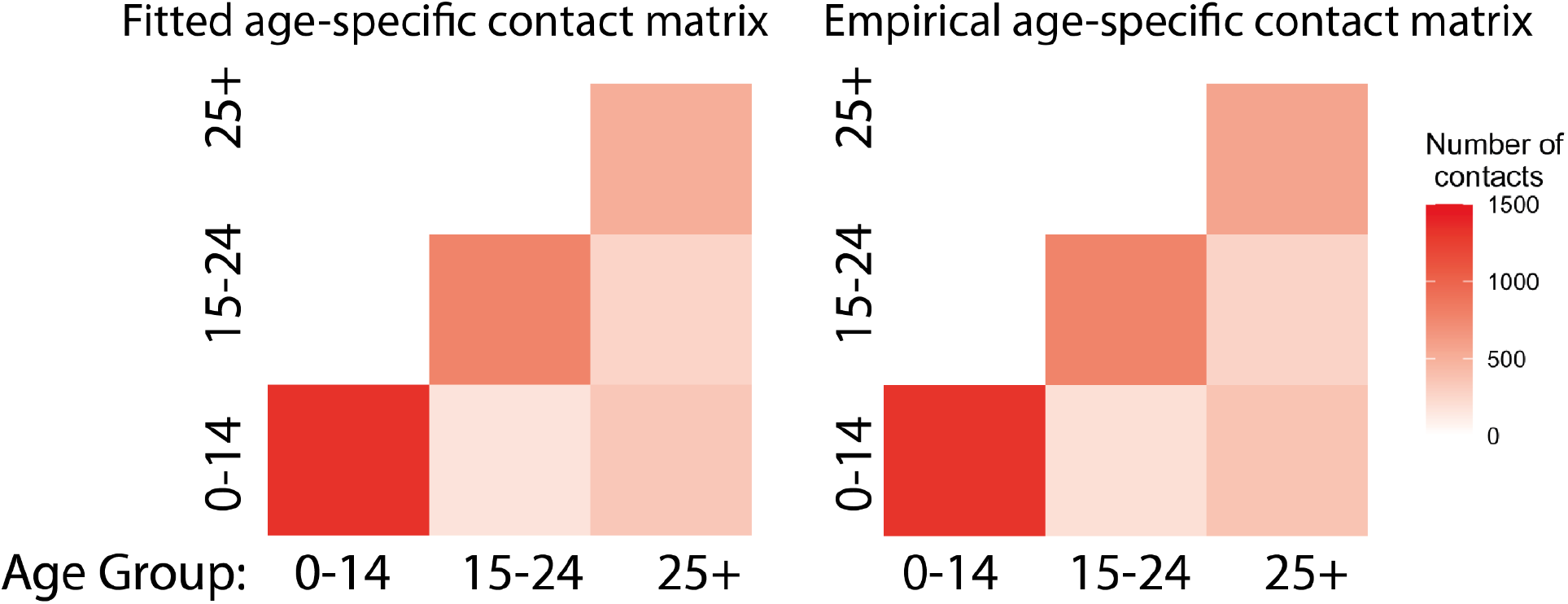
Comparison of fitted age-mixing contact matrix with those cited from survey data. The color scale shows the total number of contacts for each mixing within the populations.

**Supplementary Figure 2:**
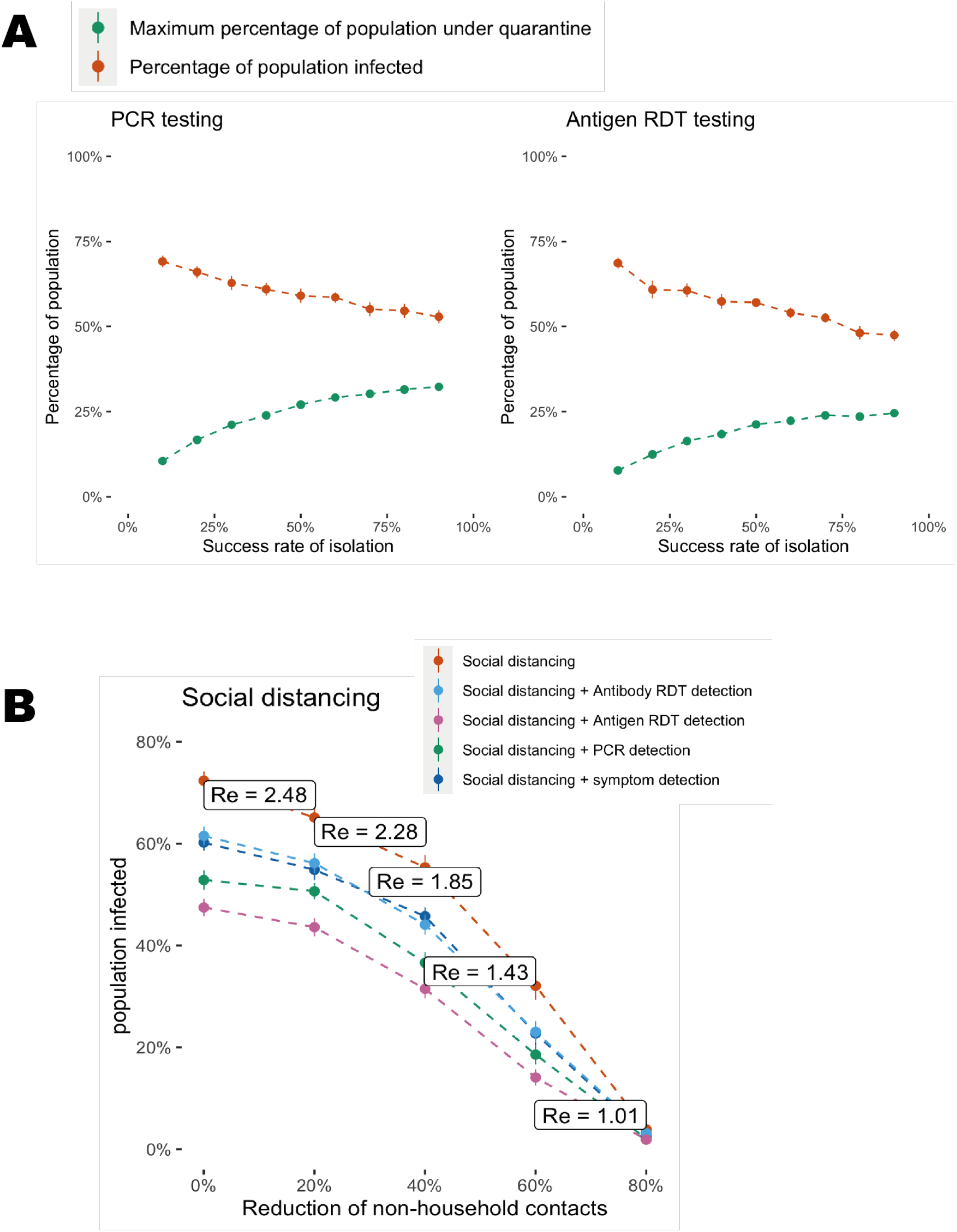
A) Proportion of populations that are infected (at the end of the trajectories) and under quarantine (max value throughout the trajectories) using PCR and antigen RDT as detection methods. The dots with bars show the mean value with 95% confidence interval from 200 simulated trajectories for each isolation compliance rate. B) Proportion of the population infected when physical distancing blocks different proportions of non-household contacts. The box shows the corresponding effective reproductive number for the level of physical distancing. The combined effect of physical distancing with each testing method shown in B is plotted with different colours.

**Supplementary Figure 3:**
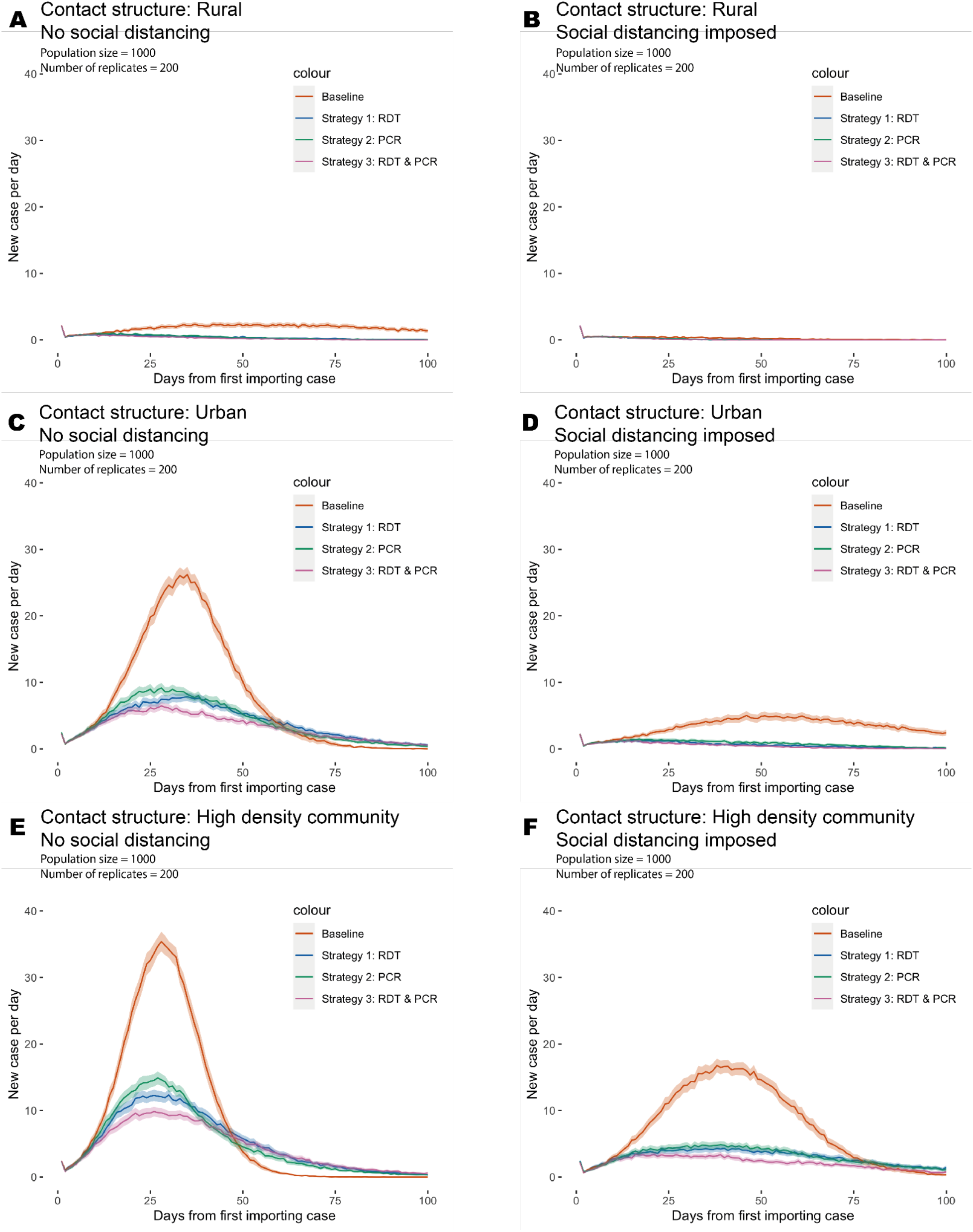
Summary of averaged outbreak trajectories in rural, urban and high density settings. A-B) Averaged trajectories of outbreaks under rural setting and those with three NPI containing strategies. Left panel shows the simulation without physical distancing and the right panel shows simulation with a physical distancing that blocks 40% of non-household contacts. C-D) Same averaged trajectories under urban setting; Left panel shows simulation without physical distancing and right panel shows simulation with a physical distancing that blocks 40% of non-household contacts. E-F) Same averaged trajectories under high density community setting; Left panel shows simulation without physical distancing and right panel shows simulation with a physical distancing that blocks 40% of non-household contacts. The curves are the mean value of daily infected numbers for a synthetic population of 1000; The shaded area indicated the 95% confidence interval of the estimation; Both mean and confidence intervals are computed from 200 simulated outbreaks.

**Supplementary Figure 4:**
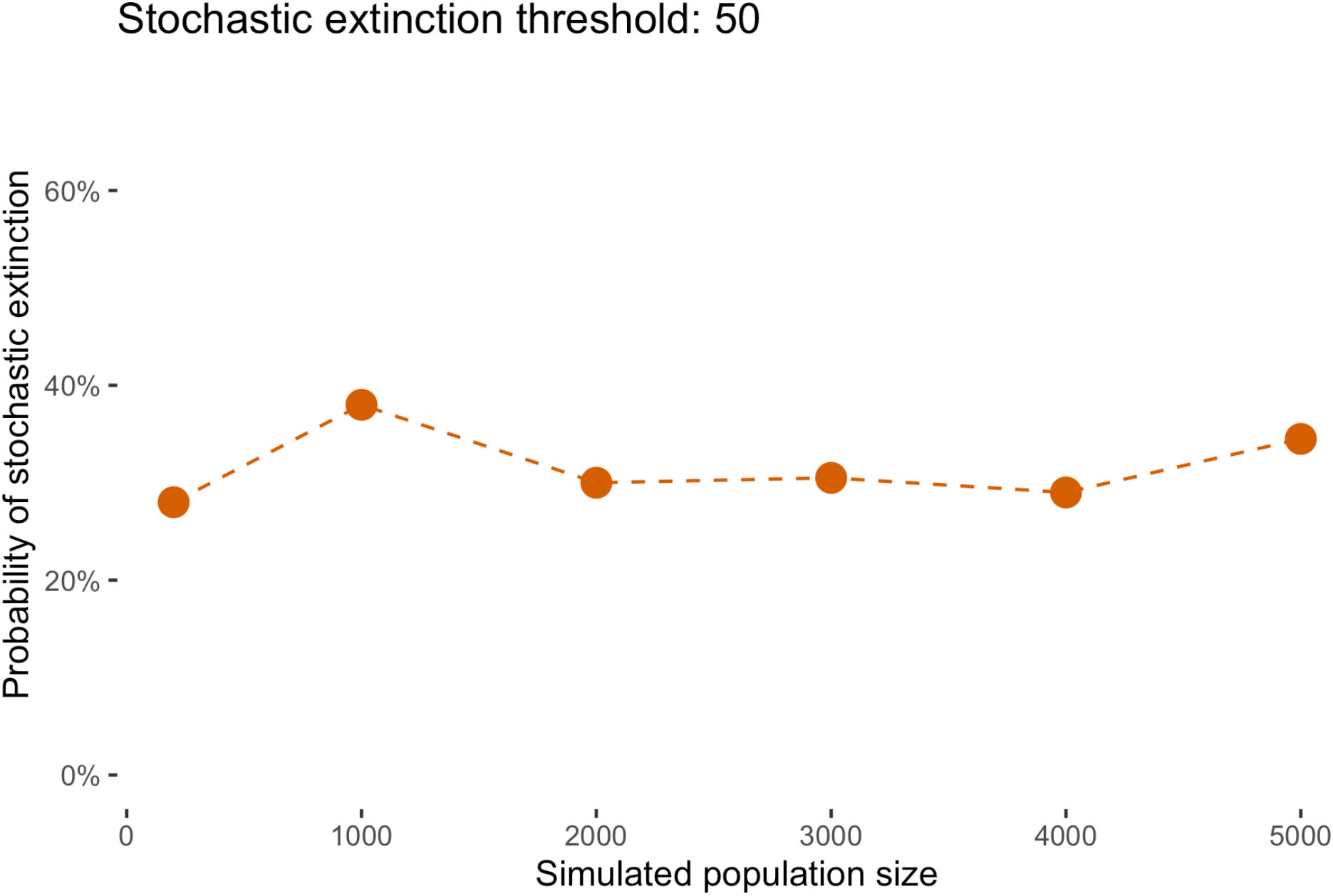
Probability of stochastic extinction against simulated population size under the rural setting. 200 bootstrap outbreaks were simulated for each population size, with stochastic extinction probability computed as the proportion of outbreaks that have less than 50 cases at the end of the 100th day.

**Supplementary Figure 5:**
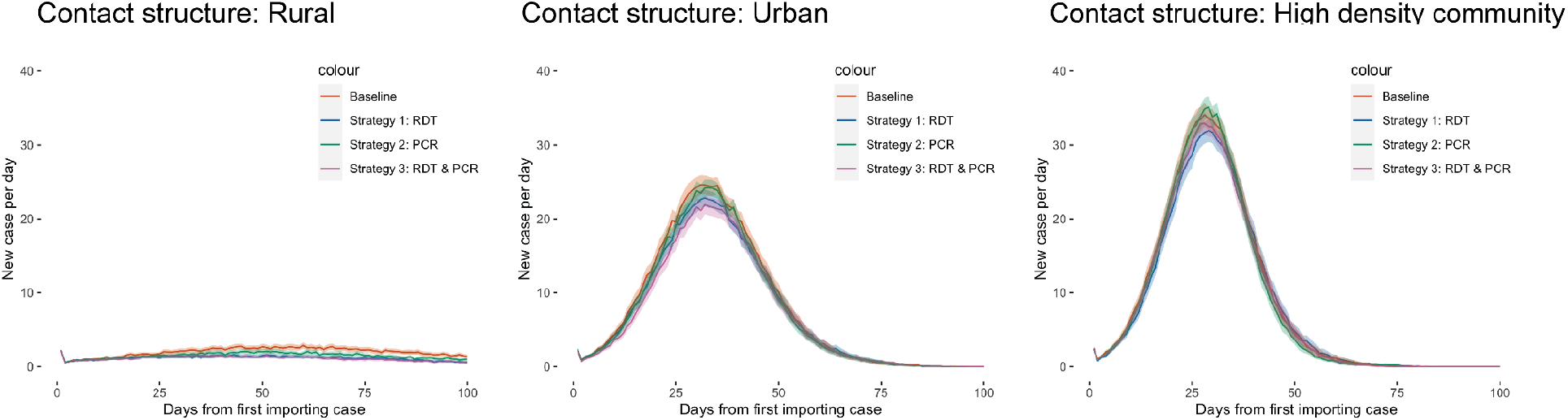
Averaged trajectories when no close contact tracing is performed under rural, urban and high density settings. The curves are the mean value of daily infected numbers for a synthetic population of 1000; The shaded area indicated the 95% confidence interval of the estimation; Both mean and confidence interval are computed from 200 simulated outbreaks.

**Supplementary Figure 6:**
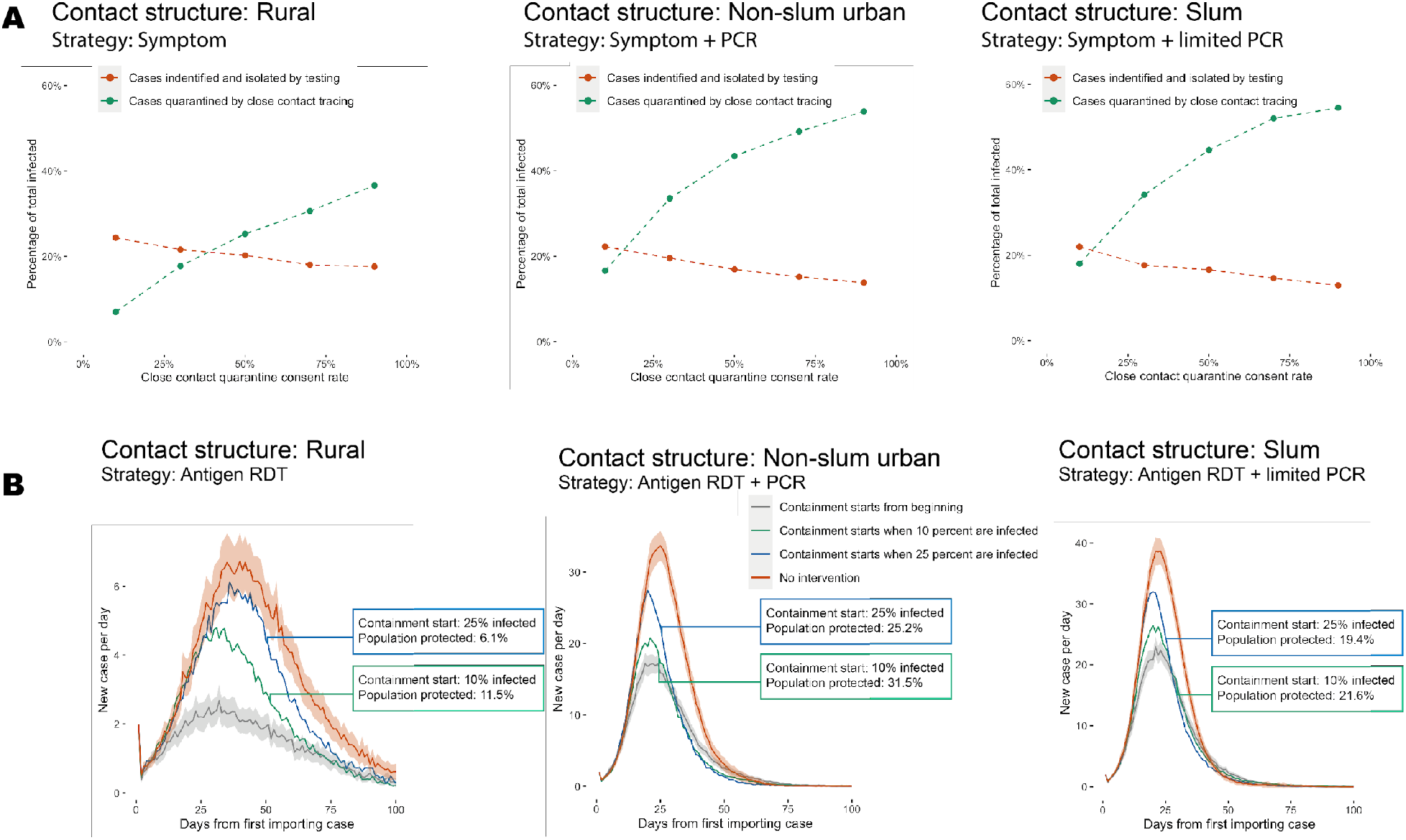
Evaluation of close contact tracing and delayed responses. A) Proportion of total infected individuals who are either isolated by testing (red dots) or quarantined by tracing (green dots) varies as the compliance rate of quarantine a close contact of a confirmed case changes. B) Implementation of containing measures after a certain proportion of the population was infected could still protect a proportion of the population, compared to the circumstances with no intervention. All simulations are performed for rural, urban and high desnity settings.

